# Diffuse myocardial fibrosis associates with incident ventricular arrhythmia in implantable cardioverter defibrillator recipients

**DOI:** 10.1101/2023.02.15.23285925

**Authors:** Eric Olausson, Jonathon Wertz, Yaron Fridman, Patrick Bering, Maren Maanja, Louise Niklasson, Timothy C Wong, Miho Fukui, João L. Cavalcante, George Cater, Peter Kellman, Syed Bukhari, Christopher A. Miller, Samir Saba, Martin Ugander, Erik B. Schelbert

## Abstract

**Background:** Diffuse myocardial fibrosis (DMF) quantified by extracellular volume (ECV) may represent a vulnerable phenotype and associate with life threatening ventricular arrhythmias more than focal myocardial fibrosis. This principle remains important because 1) risk stratification for implantable cardioverter defibrillators (ICD) remains challenging, and 2) DMF may respond to current or emerging medical therapies (reversible substrate).

**Objectives:** To evaluate the association between quantified by ECV in myocardium without focal fibrosis by late gadolinium enhancement (LGE) with time from ICD implantation to 1) appropriate shock, or 2) shock or anti-tachycardia pacing.

**Methods:** Among patients referred for cardiovascular magnetic resonance (CMR) without congenital disease, hypertrophic cardiomyopathy, or amyloidosis who received ICDs (n=215), we used Cox regression to associate ECV with incident ICD therapy.

**Results:** After a median of 2.9 (IQR 1.5-4.2) years, 25 surviving patients experienced ICD shock and 44 experienced shock or anti-tachycardia pacing. ECV ranged from 20.2% to 39.4%. No patient with ECV<25% experienced an ICD shock. ECV associated with both endpoints, e.g., hazard ratio 2.17 (95%CI 1.17-4.00) for every 5% increase in ECV, p=0.014 in a stepwise model for ICD shock adjusting for ICD indication, age, smoking, atrial fibrillation, and myocardial infarction, whereas focal fibrosis by LGE and global longitudinal strain (GLS) did not.

**Conclusions:** DMF measured by ECV associates with ventricular arrhythmias requiring ICD therapy in a dose-response fashion, even adjusting for potential confounding variables, focal fibrosis by LGE, and GLS. ECV-based risk stratification and DMF representing a therapeutic target to prevent ventricular arrhythmia warrant further investigation.

**Condensed Abstract:** Analogous to heart failure and mortality outcomes, diffuse myocardial fibrosis (DMF) quantified by extracellular volume (ECV) may represent a more vulnerable phenotype for life-threatening ventricular arrhythmia than focal myocardial fibrosis. In patients referred for cardiovascular magnetic resonance, we identified 215 subsequently receiving implantable cardioverter defibrillators (ICD). After a median of 2.9 (IQR 1.5-4.2) years, 25 patients experienced ICD shock and 44 experienced shock or anti-tachycardia pacing. ECV associated with ICD therapy in Cox regression models. Focal fibrosis variables or global longitudinal strain did not. ECV-based risk stratification and DMF representing a therapeutic target to prevent ventricular arrhythmia warrant further investigation.

## Introduction

Diffuse myocardial fibrosis (DMF) quantified by extracellular volume (ECV) may represent a vulnerable phenotype and associate with incident life threatening ventricular arrhythmias more than focal myocardial fibrosis (focal MF) detected by late gadolinium enhancement (LGE) imaging. Such relationships exist for other outcomes such as hospitalization for heart failure,(1-4) mortality(2-6), or both.(2-4,7-12) Since many implantable cardioverter defibrillators (ICD) recipients ultimately do not require their use,(13) understanding substrates for ventricular arrhythmias requiring ICD therapy remains important, especially since DMF represents a reversible substrate that may respond to current or emerging and potentially more efficacious medical therapies.(14) DMF typically affects more myocardium than focal MF given its diffuse nature. LGE from myocardial infarction (MI) or nonischemic focal MF especially involving the septum,(15,16) undoubtedly associates with ventricular arrhythmia(15-23), but most sudden cardiac death (SCD) survivors with nonischemic cardiomyopathy do not exhibit LGE.(24) Since LGE fundamentally cannot evaluate DMF,(1,2) the role of DMF in ventricular arrhythmia remains uncertain.

At the cellular level in DMF, excess myocardial collagen may impair electrical conduction and promote reentrant ventricular arrhythmia, leading to sudden cardiac death (SCD) from so-called “vulnerable myocardial interstitium”.(25) DMF may further promote ventricular arrhythmia/SCD through interactions with cardiomyocytes including decreased perfusion reserve from capillary rarefaction and perivascular fibrosis, cardiomyocyte hypoxia from increased oxygen diffusing distance, and increased afterload and preload from myocardial stiffening.(9,14) DMF occurs in both ischemic(26) and nonischemic(27) cardiomyopathy. DMF may offer a reversible substrate since it can regress significantly with 6-12 months of anti-fibrotic medical therapy blocking the renin-angiotensin-aldosterone system (RAAS).(14,28) Indeed, these agents lowered risk of sudden cardiac death (SCD) in large randomized trials. In contrast, focal MF evident on LGE persists despite treatment.(29,30)

ECV measurement using cardiovascular magnetic resonance (CMR) provides a quantitative, histologically validated, robust, and reproducible measure to quantify DMF that associates with outcomes.(31) To investigate associations between DMF and incident ventricular arrhythmia requiring ICD therapy, we studied patients referred for CMR who subsequently underwent ICD implantation. We hypothesized that DMF quantified by ECV measured in regions with no focal MF by LGE would associate with incident ventricular arrhythmias requiring ICD shock more so than other CMR stratifiers of risk, even when adjusting for potentially confounding variables. Furthermore, we hypothesized that low ECV would identify a group at especially low risk for incident arrhythmia requiring ICD shock.

## Materials and methods

### Participants

The Institutional Review Board at the University of Pittsburgh Medical Center approved the study, and all subjects provided written informed consent. All adult patients referred for contrast-enhanced CMR were recruited to participate in an observational prospective research study examining relationships between CMR data and outcomes between May 6, 2010 and March 31, 2016 at the time of CMR (n= 2,368). We excluded patients with unique disorders such as hypertrophic cardiomyopathy (n=221), cardiac amyloidosis at CMR or thereafter (n=68), or congenital heart disease (n=339). Among remaining patients, a subset of 215 received an ICD and were then followed within the UPMC integrated health network with regular ICD interrogations until November 1, 2018.

### Data Elements

Patient data were stored and managed using a REDCap database (Research Electronic Data Capture) hosted at the University of Pittsburgh. Baseline health data including demographics, comorbidity and medications were ascertained by review of medical records at the time of CMR. Ischemic cardiomyopathy was defined according to the criteria proposed by Felker et al.(32) BNP values measured in the clinical laboratory served as a summary disease severity marker, acquired at the time of CMR scanning.

Before receiving an ICD, patients underwent CMR examinations with a 1.5 Tesla scanner (Magnetom Espree; Siemens Medical Solutions) using a 32-channel phased array cardiovascular coil and dedicated CMR technologists. The examination included standard breath-held cine imaging with steady-state free precession. Left ventricular volume indices and EF were measured by experienced readers from short-axis stacks of end-diastolic and end-systolic cine frames. Global longitudinal strain (GLS) was measured with Circle cvi42 feature-tracking software (Circle Cardiovascular Imaging Inc., Calgary, Canada) from standard long axis cines as described previously.(4) LGE images using phase sensitive inversion recovery were used to identify infarcted myocardium as well as areas with focal non-ischemic scar as described previously.(33,34) Phase sensitive inversion recovery LGE prevented artifacts from short inversion times that can mimic midwall fibrosis. The extent of focal fibrosis by LGE was assessed visually in terms of the extent of LGE (none, <25%, 26% to 50%, 51% to 75%, >75%), rendering 5 categories for each of the 17 segments to compute approximate extent of LGE expressed as a percentage of left ventricular mass. Clinicians caring for the patient had access to all CMR data prior to ICD placement except for ECV data.

### Quantification of DMF with ECV

We employed reproducible(35) and validated(14) ECV measures after an intravenous bolus of a gadoteridol (0.2 mmol/kg, Prohance, Bracco Diagnostics, Princeton, NJ) as described previously (i.e., Modified Look Locker Inversion recovery, 5 and 2 image sampling scheme following 2 inversion pulses pre-contrast, 4-3-2 sampling scheme following 3 inversion pulses post contrast).(5) To focus exclusively on DMF, native T1 and ECV measurement occurred only in regions completely free of focal LGE, whether MI or nonischemic focal MF, using the clinical CMR report as the arbiter of what constituted significant LGE. We measured T1 data in the middle third of the myocardial wall to avoid partial volume effects, avoiding voxels stratifying the border between blood pool and myocardium.

We quantified DMF with ECV(36,37) defined as:

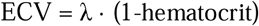

where λ = [ΔR1_myocardium_] / [ΔR1_bloodpool_] pre and post gadolinium contrast (where R1=1/T1) from basal and mid-ventricular short axis slices in noninfarcted myocardium as described previously.(1,5,35,38) Hematocrit measures were acquired on the day of scanning and measured in the clinical laboratory. We measured ECV blinded to all ICD and comorbidity data. For comparison purposes, we also report ECV values that included myocardium with nonischemic focal myocardial fibrosis atypical of myocardial infarction as per a previously used convention.(2)

### Follow-up and outcomes

The primary end-point included an episode of ventricular arrhythmia requiring ICD shock according to parameters chosen by the board-certified clinical electrophysiologist responsible for the patient’s care. The secondary end-point included episodes of ventricular arrhythmia requiring either ICD shock or anti-tachycardia pacing (ATP). Patients had their ICD devices interrogated at regular intervals after implantation, either during clinical visits or via remote interrogation. To ensure the validity of the endpoints, the primary arrhythmia recordings requiring ICD therapy were independently reviewed and adjudicated by two experienced electrophysiologists (SS and JW) blinded to ECV data and all other clinical data. Discrepant interpretation for five ICD therapy episodes warranted a second joint adjudication to reach consensus.

### Statistical analysis

We summarized categorical variables as numbers and percentages and continuous variables as medians and interquartile range (IQR) since some showed skewed non-normal distributions based on the Kolmogorov-Smirnov test. Chi square (χ^2^) tests or Fisher’s exact test compared categorical variables, and nonparametric Wilcoxon rank sum tests compared continuous variables according to whether the primary outcome of ICD shock occurred. Survival analysis examined time to events commencing with ICD placement, not CMR, to minimize potential for ascertainment bias since ICD implantation denotes clinical assessment of sizable SCD risk and permits arrhythmia detection. Survival analysis was limited to survivors only by right censoring for death. The log-rank test and Cox regression examined time until (1) first ICD shock, and (2) first ICD shock or ATP, with the latter endpoint with more frequent events serving as a secondary analysis. Non-significant time interaction terms for ECV (i.e., product of ECV and follow-up time) confirmed the proportional hazard assumption in Cox regression models.

In Cox regression models, we expressed ECV as a continuous variable (percentage) and reported hazard ratios (HR) scaled as 5% increments to scale the HR to generate clinically meaningful intervals. Similarly, all continuous variables in regression models were modeled as such but scaled to clinically meaningful intervals. To benchmark outcome associations between ECV against other clinically important variables in Cox regression models, we compared their chi square (χ^2^) values which remain constant regardless of how one chooses to scale the HR. We also examined native T1 (measured in the same area as ECV) which is weaker measure of DMF that does not require contrast and is not specific for the myocardial interstitium where DMF occurs.(39)

In multivariable Cox regression models, we tested for interactions between ECV and other clinically relevant variables by including a term that was the product of the paired variables: age, sex, EF, presence of infarction by LGE, septal midwall myocardial fibrosis by LGE, or any non-ischemic scar by LGE. Patients were censored when reaching the endpoint or at the time of last ICD interrogation. Given limited statistical power due to limited events, we created two parsimonious multivariable models, one “clinical” model leveraging clinical knowledge and another model using automated stepwise selection. Given limited events and the rule of thumb employing one predictor variable per 5-10 events to prevent model over fitting, we stratified by ICD indication (primary or secondary) to adjust for this variable while conserving degrees of freedom.

We created multiple Cox regression models to ensure consistent results. The “clinical” model selected covariates informed by clinical acumen. The clinical model for ICD shock only in Table 2 stratified for ICD indication (primary versus secondary prevention) and included ECV, log BNP, extent of MI, extent of nonischemic LGE as covariates. The clinical model for ICD shock or ATP therapies in Table 3 was identical but added diabetes and coronary artery bypass surgery as covariates. The “stepwise” models simply selected covariates based on strength of associations with outcomes which required a p value of <0.10 to enter and remain in the model. Statistical tests were two-sided, and p-values <0.05 were considered significant. Statistical analyses were performed using SAS 9.3 (SAS Institute, Cary, NC).

**Table 1.**
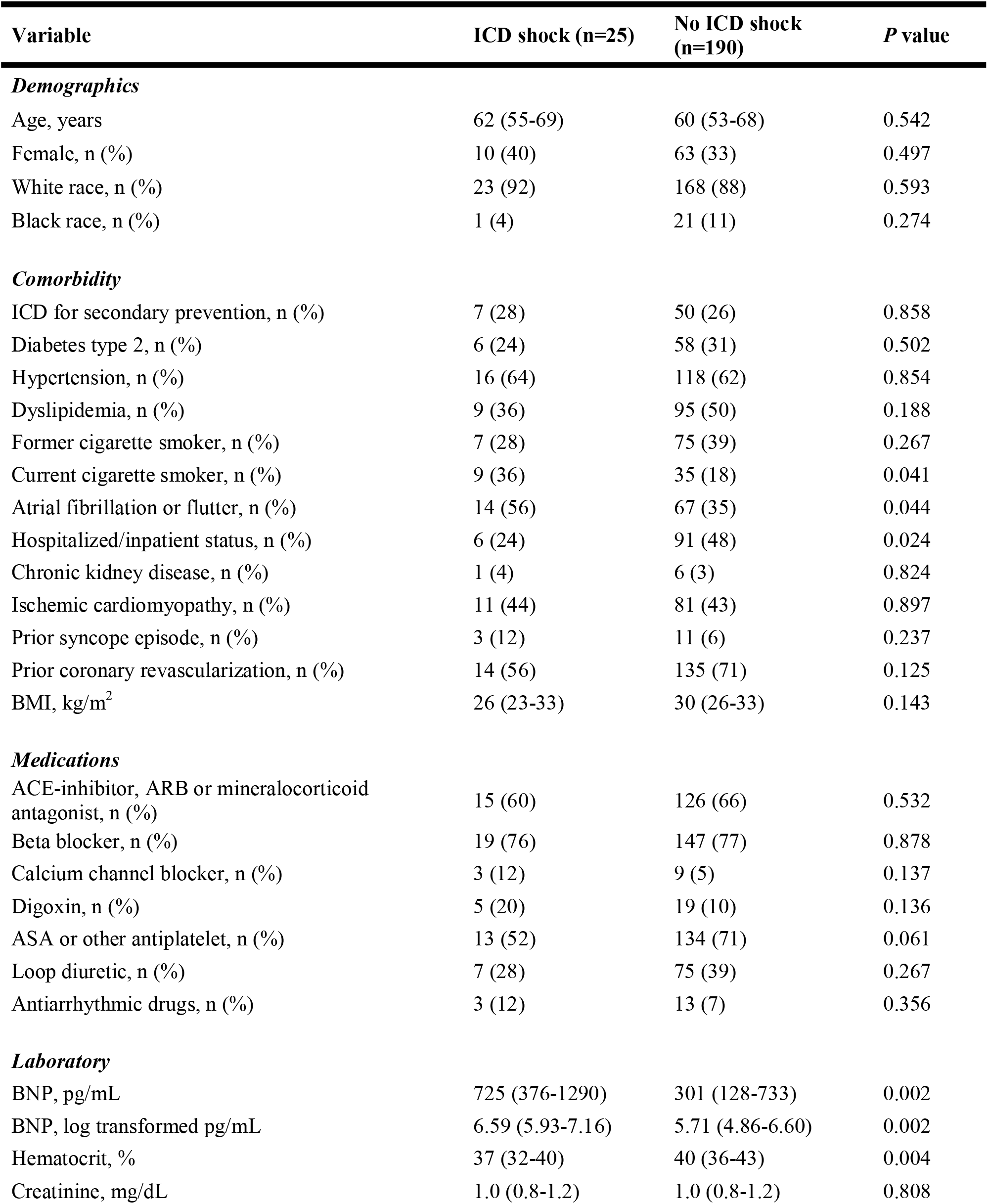
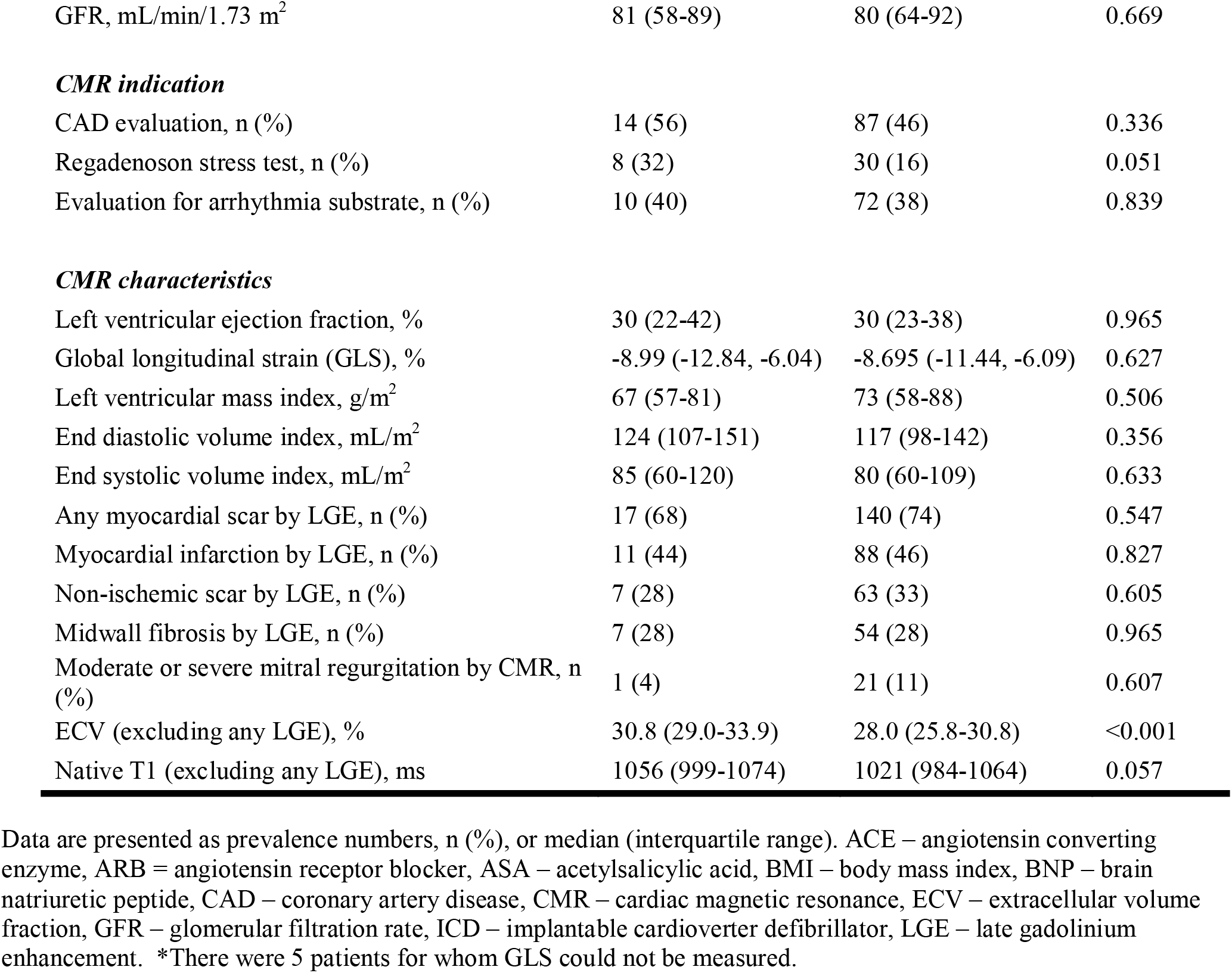
Baseline patient characteristics (n=215) at time of CMR according to whether patient experienced arrhythmia requiring intervention in form of ICD shock during the follow up period.

| Variable | ICD shock (n=25) | No ICD shock (n=190) | P value |
| --- | --- | --- | --- |
| ***Demographics*** |  |  |  |
| Age, years | 62 (55-69) | 60 (53-68) | 0.542 |
| Female, n (%) | 10 (40) | 63 (33) | 0.497 |
| White race, n (%) | 23 (92) | 168 (88) | 0.593 |
| Black race, n (%) | 1 (4) | 21 (11) | 0.274 |
| ***Comorbidity*** |  |  |  |
| ICD for secondary prevention, n (%) | 7 (28) | 50 (26) | 0.858 |
| Diabetes type 2, n (%) | 6 (24) | 58 (31) | 0.502 |
| Hypertension, n (%) | 16 (64) | 118 (62) | 0.854 |
| Dyslipidemia, n (%) | 9 (36) | 95 (50) | 0.188 |
| Former cigarette smoker, n (%) | 7 (28) | 75 (39) | 0.267 |
| Current cigarette smoker, n (%) | 9 (36) | 35 (18) | 0.041 |
| Atrial fibrillation or flutter, n (%) | 14 (56) | 67 (35) | 0.044 |
| Hospitalized/inpatient status, n (%) | 6 (24) | 91 (48) | 0.024 |
| Chronic kidney disease, n (%) | 1 (4) | 6 (3) | 0.824 |
| Ischemic cardiomyopathy, n (%) | 11 (44) | 81 (43) | 0.897 |
| Prior syncope episode, n (%) | 3 (12) | 11 (6) | 0.237 |
| Prior coronary revascularization, n (%) | 14 (56) | 135 (71) | 0.125 |
| BMI, kg/m<sup>2</sup> | 26 (23-33) | 30 (26-33) | 0.143 |
| ***Medications*** |  |  |  |
| ACE-inhibitor, ARB or mineralocorticoid antagonist, n (%) | 15 (60) | 126 (66) | 0.532 |
| Beta blocker, n (%) | 19 (76) | 147 (77) | 0.878 |
| Calcium channel blocker, n (%) | 3 (12) | 9 (5) | 0.137 |
| Digoxin, n (%) | 5 (20) | 19 (10) | 0.136 |
| ASA or other antiplatelet, n (%) | 13 (52) | 134 (71) | 0.061 |
| Loop diuretic, n (%) | 7 (28) | 75 (39) | 0.267 |
| Antiarrhythmic drugs, n (%) | 3 (12) | 13 (7) | 0.356 |
| ***Laboratory*** |  |  |  |
| BNP, pg/mL | 725 (376-1290) | 301 (128-733) | 0.002 |
| BNP, log transformed pg/mL | 6.59 (5.93-7.16) | 5.71 (4.86-6.60) | 0.002<sup>23</sup> |
| Hematocrit, % | 37 (32-40) | 40 (36-43) | 0.004 |
| Creatinine, mg/dL | 1.0 (0.8-1.2) | 1.0 (0.8-1.2) | 0.808 |
| GFR, mL/min/1.73 m<sup>2</sup> | 81 (58-89) | 80 (64-92) | 0.669 |
| ***CMR indication*** |  |  |  |
| CAD evaluation, n (%) | 14 (56) | 87 (46) | 0.336 |
| Regadenoson stress test, n (%) | 8 (32) | 30 (16) | 0.051 |
| Evaluation for arrhythmia substrate, n (%) | 10 (40) | 72 (38) | 0.839 |
| ***CMR characteristics*** |  |  |  |
| Left ventricular ejection fraction, % | 30 (22-42) | 30 (23-38) | 0.965 |
| Global longitudinal strain (GLS), % | -8.99 (-12.84, -6.04) | -8.695 (-11.44, -6.09) | 0.627 |
| Left ventricular mass index, g/m<sup>2</sup> | 67 (57-81) | 73 (58-88) | 0.506 |
| End diastolic volume index, mL/m<sup>2</sup> | 124 (107-151) | 117 (98-142) | 0.356 |
| End systolic volume index, mL/m<sup>2</sup> | 85 (60-120) | 80 (60-109) | 0.633 |
| Any myocardial scar by LGE, n (%) | 17 (68) | 140 (74) | 0.547 |
| Myocardial infarction by LGE, n (%) | 11 (44) | 88 (46) | 0.827 |
| Non-ischemic scar by LGE, n (%) | 7 (28) | 63 (33) | 0.605 |
| Midwall fibrosis by LGE, n (%) | 7 (28) | 54 (28) | 0.965 |
| Moderate or severe mitral regurgitation by CMR, n (%) | 1 (4) | 21 (11) | 0.607 |
| ECV (excluding any LGE), % | 30.8 (29.0-33.9) | 28.0 (25.8-30.8) | <0.001 |
| Native T1 (excluding any LGE), ms | 1056 (999-1074) | 1021 (984-1064) | 0.057 |

Data are presented as prevalence numbers, n (%), or median (interquartile range). ACE – angiotensin converting enzyme, ARB = angiotensin receptor blocker, ASA – acetylsalicylic acid, BMI – body mass index, BNP – brain natriuretic peptide, CAD – coronary artery disease, CMR – cardiac magnetic resonance, ECV – extracellular volume fraction, GFR – glomerular filtration rate, ICD – implantable cardioverter defibrillator, LGE – late gadolinium enhancement. \*There were 5 patients for whom GLS could not be measured.

**Table 2.**
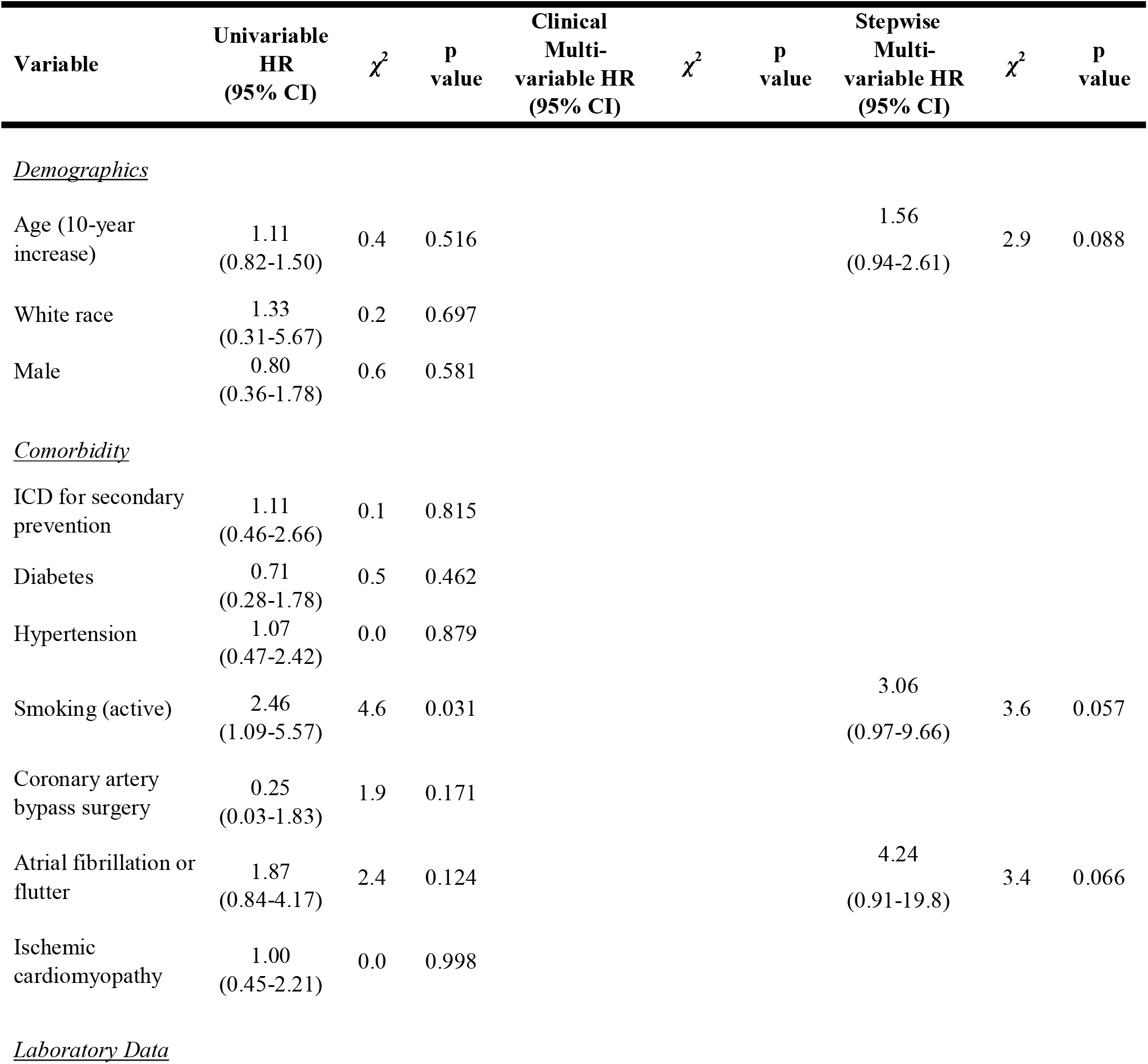
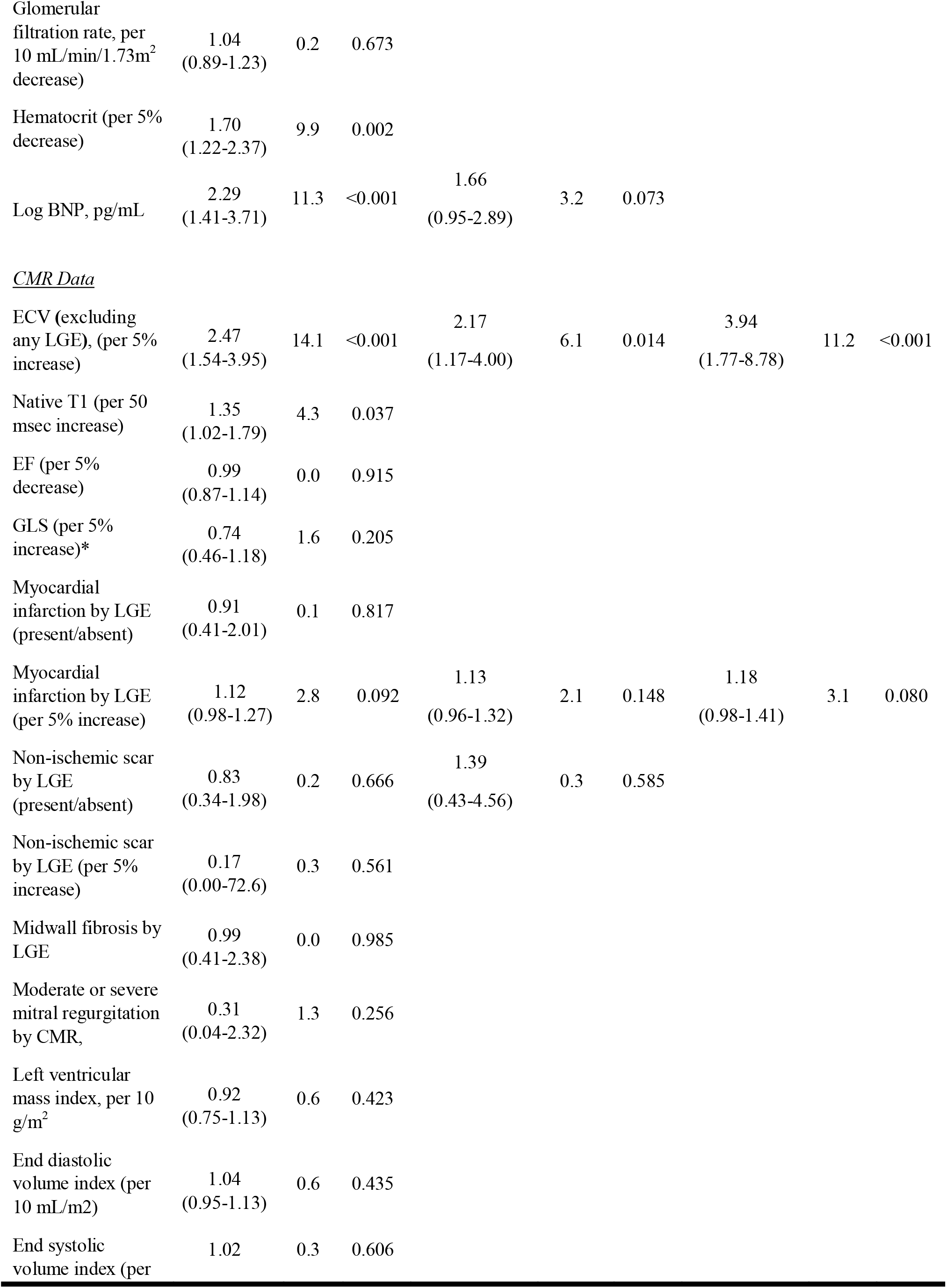
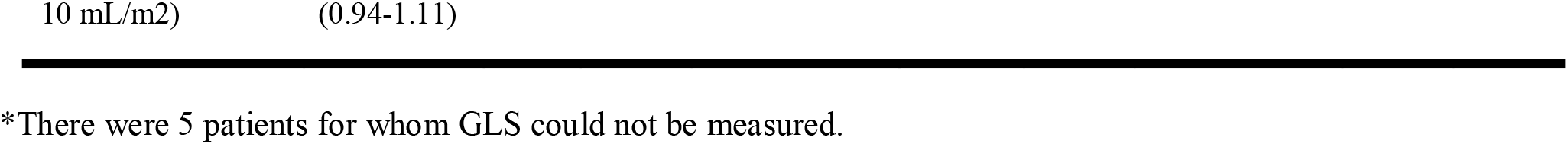
Univariable and multivariable Cox regression models demonstrated associations between incident ICD shock (N=25) and diffuse myocardial fibrosis measured by ECV in myocardium without LGE. The multivariable models stratified for ICD indication (primary versus secondary prevention). The clinical model adjusted for variables believed to represent principal mediators of risk on clinical grounds (i.e., ECV, BNP, extent of myocardial infarction, and extent of focal myocardial fibrosis) within the constraints of limited numbers of events. The stepwise model employed automatic selection of variables associated with outcomes (based on p<0.10) stratified by ICD indication.

**Table 3.**
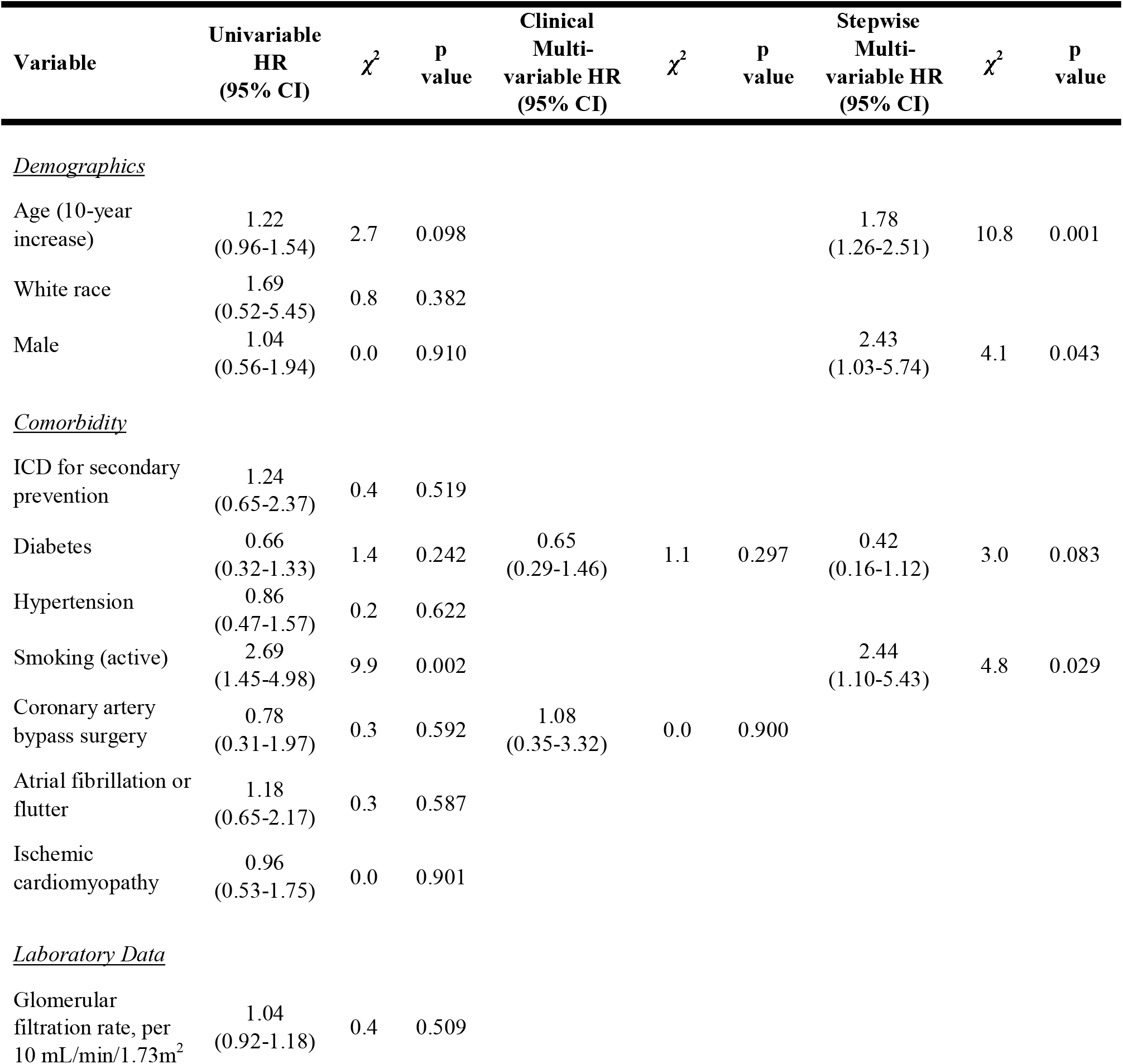
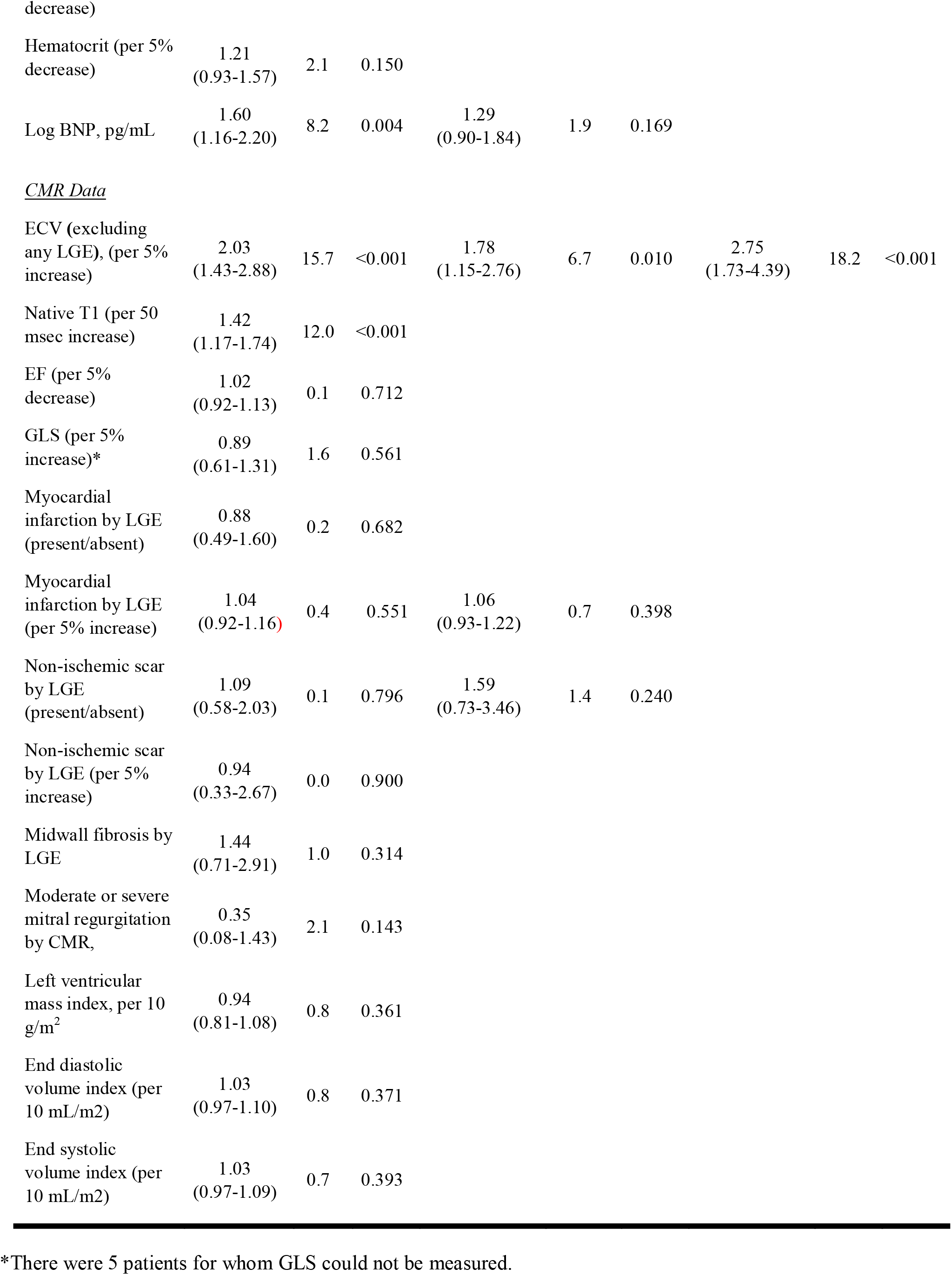
For the composite endpoint of either incident ICD shock or incident ATP therapy (N=44), univariable and multivariable Cox regression models demonstrated associations with diffuse myocardial fibrosis measured by ECV (in myocardium without LGE). The multivariable models stratified for ICD indication (primary versus secondary prevention). The clinical model adjusted for variables believed to represent principal mediators of risk on clinical grounds (e.g., BNP, myocardial infarction, and focal myocardial fibrosis) within the constraints of limited numbers of events. The stepwise model employed automatic selection of variables associated with outcomes (where p<0.10) stratified by ICD indication.

## Results

### Patient characteristics

The final study cohort included 215 patients. ICDs were implanted across 5 hospitals within the UPMC health system. Most ICD recipients were older with low ejection fraction and a high prevalence of LGE. ECV ranged from 20.2% to 39.4%. Baseline characteristics are summarized in Table 1 according to whether individuals subsequently experienced the primary end-point of ICD shock. The two groups appeared similar clinically, except that individuals experiencing ICD shock were more likely to smoke, be hospitalized at the time of CMR, have atrial fibrillation, have higher levels of brain natriuretic peptide (BNP), and have a lower hematocrit. Median time between CMR and subsequent ICD implantation was 36 (IQR 3-147) days.

Patients who experienced ICD shocks had higher ECV (p<0.001). There were 18 inappropriate shocks, but ECV did not differ in those with or without inappropriate shocks (median ECV 28.2% for both, p=0.662). The prevalence of midwall focal fibrosis or even any focal fibrosis involving the interventricular septum (a less restrictive definition) did not differ according to whether patients experienced ICD shock. In fact, no other metric of myocardial damage exhibited differences according to incident ICD shock (Table 1). Patients who experienced ICD shocks also trended towards having a higher native T1 (p=0.057).

### Associations between ECV and incident ventricular arrhythmias

During a median follow-up of 2.9 (IQR 1.5-4.2) years after ICD implantation, 25 (12%) patients developed ventricular arrhythmias requiring termination by ICD shock while 44 (20%) patients had episodes requiring either ICD shock or ATP therapy, including 5 episodes of ventricular fibrillation. Median cycle length triggering therapy was 299 msec (IQR 250-330 msec). ECV did not interact significantly with age, sex, EF, presence of infarction, septal midwall myocardial fibrosis, or any non-ischemic scar. Kaplan-Meier plots demonstrated that ECV measures of DMF associated with both end-points in a dose-response fashion whereby higher ECV category associated with higher risk of 1) ICD shock, and 2) either ICD shock or ATP (p<0.001 for all, Figure 1). Notably, no patients with ECV<25% (n=38, 17%) experienced ICD shocks. In contrast, midwall focal fibrosis involving the interventricular septum did not exhibit any associations with outcomes (Figure 2). Native T1 measures of DMF exhibited less consistent associations overall (Supplemental Figure).

**Figure 1.**
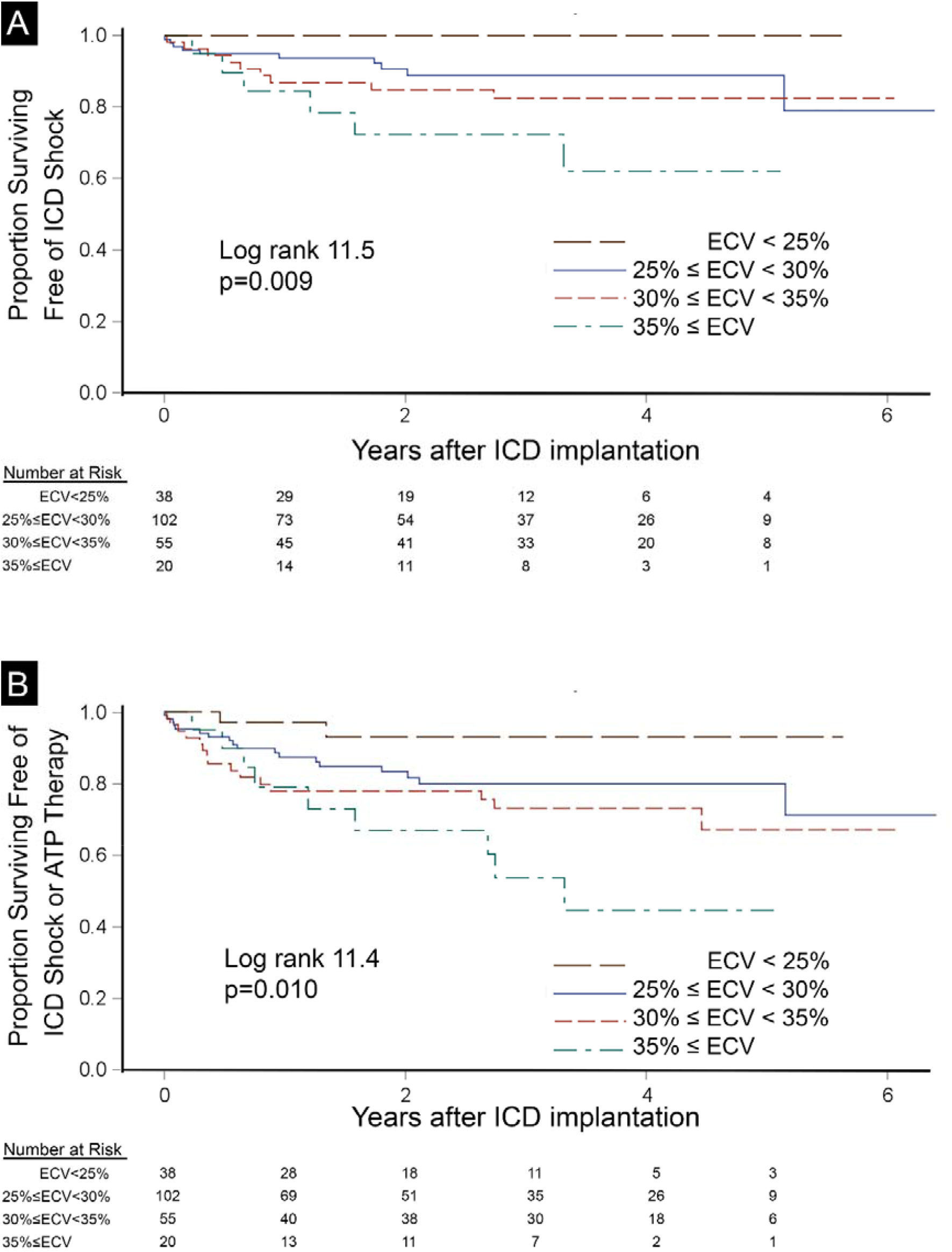
Diffuse myocardial fibrosis measured by ECV associated with incident ICD shock (n=25, panel A) or the composite endpoint of ICD shock or ATP therapy (n=44, panel B) in 215 ICD recipients exhibiting a dose response fashion. These associations remained in all multivariable models.

**Figure 2.**
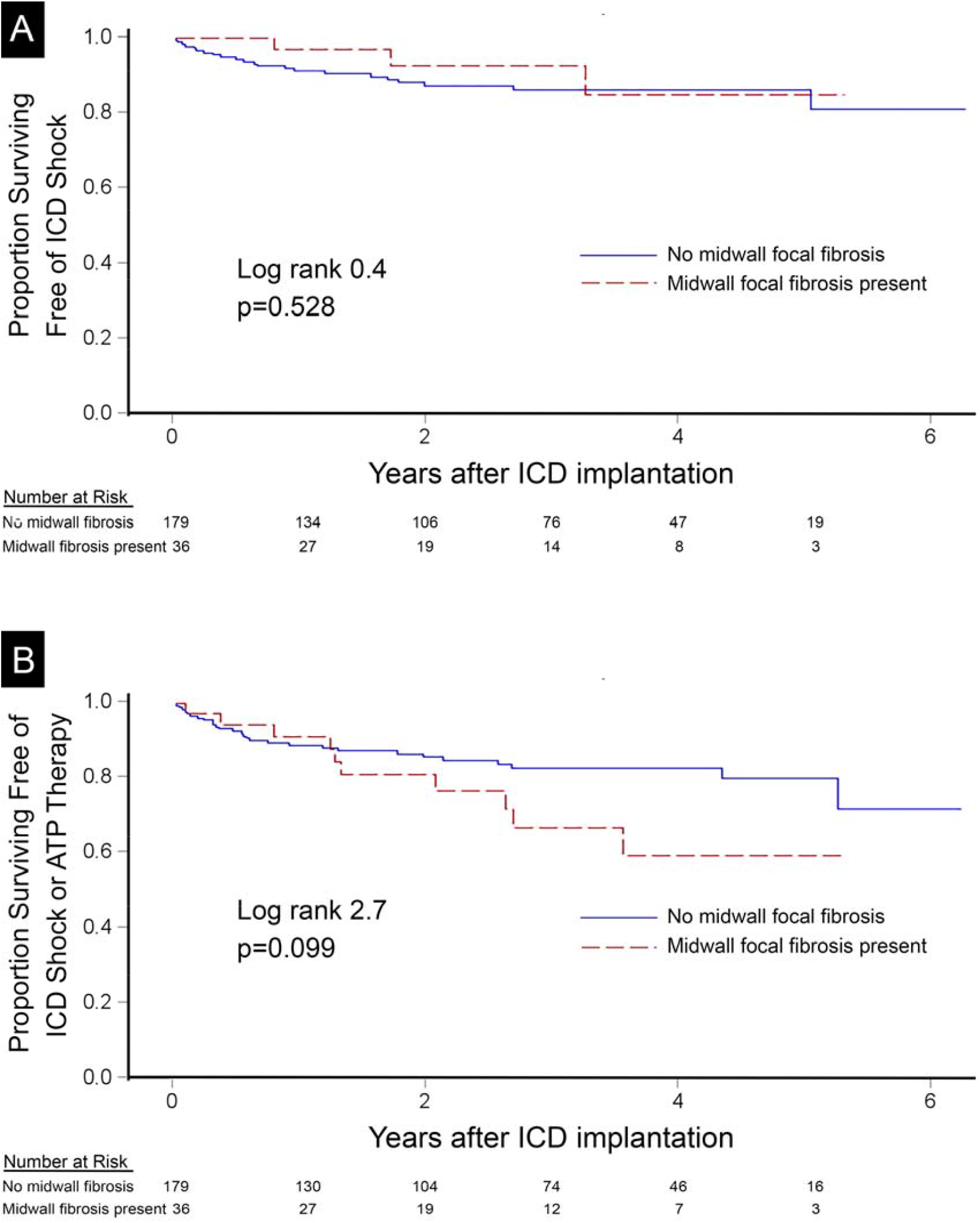
Midwall focal fibrosis by LGE-CMR in the interventricular septum did not associate with incident ICD shock (n=25, panel A) or the composite endpoint of ICD shock or ATP therapy (n=44, panel B) in 215 ICD recipients.

In univariable Cox regression models, ECV associated more strongly with 1) ICD shock or 2) either ICD shock or ATP compared to midwall focal fibrosis in the interventricular septum, myocardial infarction, any nonischemic focal fibrosis, or native T1, as shown by the χ^2^ data (Tables 2 and 3). Ignoring issues of collinearity, when ECV and native T1 were both combined in a Cox model, only ECV associated with ICD shock (HR 2.3, 95%CI 1.40-3.84 per 5% ECV increase, χ^2^=10.6, p=0.001 vs. HR 1.12, 95%CI 0.82-1.53 per 50 msec native T1 increase, χ^2^=0.5, p=0.485, respectively), whereas both associated with either ICD shock or ATP but ECV exhibited stronger associations (HR 1.78, 95%CI 1.22-2.60 per 5% increase, χ^2^=8.8, p=0.003 vs. HR 1.25, 95%CI 1.01-1.56 per 50 msec native T1 increase, χ^2^=4.1, p=0.044, respectively).

In multivariable Cox regression models, ECV expressed as a continuous variable associated with incident 1) ICD shock or 2) the secondary endpoint of either ICD shock or ATP even after adjusting for other potential variables that may predispose to ventricular arrhythmias such as: age, EF, GLS, presence of infarction or non-ischemic scar on LGE-CMR, ICD indication (primary or secondary prevention) and diagnosis of ischemic cardiomyopathy. Myocardial infarction (expressed as a binary or continuous variable (% left ventricular mass), septal midwall focal myocardial fibrosis, any nonischemic scar (expressed as a binary or continuous variable) did not associate with incident 1) ICD shock or 2) the secondary endpoint of either ICD shock or ATP. GLS also did not associate with ICD therapy. BNP, hypertension, and hematocrit did not associate with outcomes sufficiently to enter either of the stepwise multivariable models using the p=0.10 threshold. Tables 2 and 3 summarize the Cox regression data for these endpoints.

## Discussion

In this cohort of surviving patients who underwent CMR and subsequent ICD implantation, we demonstrate that ECV measures of DMF associated robustly with incident ventricular arrhythmias requiring 1) ICD shock, or 2) ICD shock or ATP therapy in a dose-response fashion. Similar relationships exist between ECV and other outcomes such as hospitalization for heart failure,(1-4) mortality(2-6), or both.(2-4,7-12) Furthermore, ECV measures of DMF associated with incident arrhythmia more so than focal myocardial fibrosis detected by LGE, whether myocardial infarction, nonischemic midwall myocardial fibrosis in the septum or elsewhere. Native T1 measures of DMF which do not require contrast and GLS did not exhibit robust associations with ICD therapies. Associations between ECV and ICD therapy remained significant even when controlling for other important conditions in various multivariable models, including ischemic cardiomyopathy and primary/secondary prevention indication. Finally, those with minimal DMF, i.e., ECV<25%, had 100% negative predictive value for the primary outcome with no incident events over the study period, acknowledging limited sample size and limited follow-up.

Several data support ECV-based risk stratification for arrhythmia. Analogous to DMF, the precedent of “vulnerable interstitium” from cardiac amyloidosis with a high prevalence of ventricular arrhythmia requiring ICD therapy illustrates how diffuse protein (amyloid) deposition in the myocardial interstitium may distort microarchitecture and predispose to incident ventricular arrhythmia,(40) similar to excess collagen protein.(25) DMF promotes arrhythmia whereby excess collagen between cardiomyocytes impairs electrical conduction and creates a substrate for reentry. We postulate that diffuse fibrosis being dispersed widely throughout the myocardium probably jeopardizes more total myocardium and thus promotes arrhythmia more than focal myocardial fibrosis which typically involves only small portions of myocardium. A recent smaller study modeling only 11 arrhythmic events in patients with nonischemic cardiomyopathy over a median follow-up of 21 months reported similar results.(41)

Our results and those reported by Di Marco and colleagues(41) differ from others who reported significant associations with ICD therapies only for native T1, not ECV.(42,43) Reasons underlying these differences remain uncertain, but we consider these issues: We note that our sample was somewhat larger, with longer follow-up, and more ICD events, and we sampled more of the left ventricular myocardium to minimize sample error. We also used a different scanner vendor that employed T1 mapping leveraging motion correction technology and phase sensitive reconstruction.(44,45) The robustness of T1 measurement may vary by vendor. Understanding vendor differences for T1 mapping requires further investigation.

Our results also differ from several prior reports associating LGE with life-threatening ventricular arrhythmia. We suspect the lack of association between LGE and ICD therapies in our cohort simply reflects limited statistical power related to limited sample size and follow-up. We note remarkably similar prevalences of various LGE patterns in our cohort that align with the works of others (Supplemental Table).

Given the observation that substantial proportions of ICD recipients never require ICD therapies, and given their costs and their risks including infection and inappropriate shock, optimal risk stratification requires further understanding, especially for primary prevention in nonischemic cardiomyopathy as exemplified by the DANISH Trial.(46) The significant proportion of SCD survivors who do not exhibit focal MF with LGE(24) underscores the challenges further. Whether more robust phenotyping provided by ECV quantification of DMF improves risk-stratification and identifies high risk subgroups with DMF ultimately requires randomized trials of ECV-guided care to establish benefit (analogous to LGE guided-care under investigation, NCT05568069, NCT01918215).(47) Similarly, whether reversal of DMF with antifibrotic therapy lowers risks of incident ventricular arrhythmia and SCD(48) requires clinical trials, especially with anti-fibrotic medications more efficacious than the modestly effective agents currently available.(14,41)

Our study has several limitations. First, sample size was limited which may introduce type 2 statistical error and reduce power to detect established associations between arrhythmia and hypertension,(25) LGE,(15-23) T1,(42,44,45) or GLS,(49) and the sample was constrained to a single center which may limit generalizability. Despite the limited sample size and limited follow-up duration, we demonstrated novel dose-response relationships suggesting ECV measures of DMF associates with incident ICD therapies more robustly than other LGE, T1, or GLS phenotypes. Second, observational data may not control for residual confounding. Still, we controlled for several clinically relevant variables including EF, ischemic heart disease, and ICD indication. Third, observational data do not establish causality. Whether antifibrotic therapy lowers incident ventricular arrhythmia and SCD requires further study with randomized controlled trials, and our work supports such trials. Fourth, programmed zones for delivering ICD therapies may vary by electrophysiologist which can influence the results, and some arrhythmias may have self-terminated, thus inflating associations. Nonetheless, our data reflect the conventional practice of multiple board-certified electrophysiologists serving 5 hospitals. Fifth, we did not apply thresholding techniques to quantify LGE extent, but recent work shows marginal value for LGE quantification of nonischemic fibrosis beyond expressing LGE as a binary variable.(50) Finally, we lacked histological validation of our ECV measures, but others have repeatedly validated ECV previously which likely represents the most robust noninvasive measure available.(2,14) Transient myocardial edema and inflammation may increase both ECV and native T1, but only ECV measures yielded robust risk stratification suggesting that diffuse myocardial fibrosis underlies the associations between ECV and incident arrhythmia occurring long after the baseline CMR scan.

## Conclusions

ECV measures of DMF associate with incident arrhythmia requiring ICD therapy in a dose-response fashion. ECV associates with incident arrhythmia more so than focal myocardial fibrosis detected by LGE (e.g., myocardial infarction, septal midwall focal MF, or nonischemic myocardial scar elsewhere). Native T1 measures of DMF and GLS did not exhibit robust associations. Patients without DMF as measured by ECV appear to have a very low short-term risk of ventricular arrhythmia. The ability of ECV to stratify risk of incident ventricular arrhythmia and SCD and the suitability of DMF as a therapeutic target with efficacious medication to lower these risks both warrant further investigation.

## Data Availability

All data produced in the present study may be available upon reasonable request to the authors

## Abbreviations

ATP: anti-tachycardia pacing
CMR: cardiovascular magnetic resonance
DMF: diffuse myocardial fibrosis
ECV: extracellular volume
Focal MF: focal myocardial fibrosis
GLS: global longitudinal strain
ICD: implantable cardioverter defibrillator
LGE: late gadolinium enhancement
SCD: sudden cardiac death

## Clinical Perspectives

### Clinical competencies

ECV measures of diffuse myocardial fibrosis stratifies risk of requiring ICD therapies where the higher the burden of DMF, the higher the risks of requiring ICD therapies to abort an ostensible life threatening arrhythmia. ECV may help identify patients at risk who would benefit from ICD placement.

### Translational outlook

The ability of ECV to stratify risk of incident ventricular arrhythmia and SCD, e.g., ECV guided care, and the suitability of DMF as a therapeutic target to lower risk both warrant further investigation.

---

**Central Illustration.**
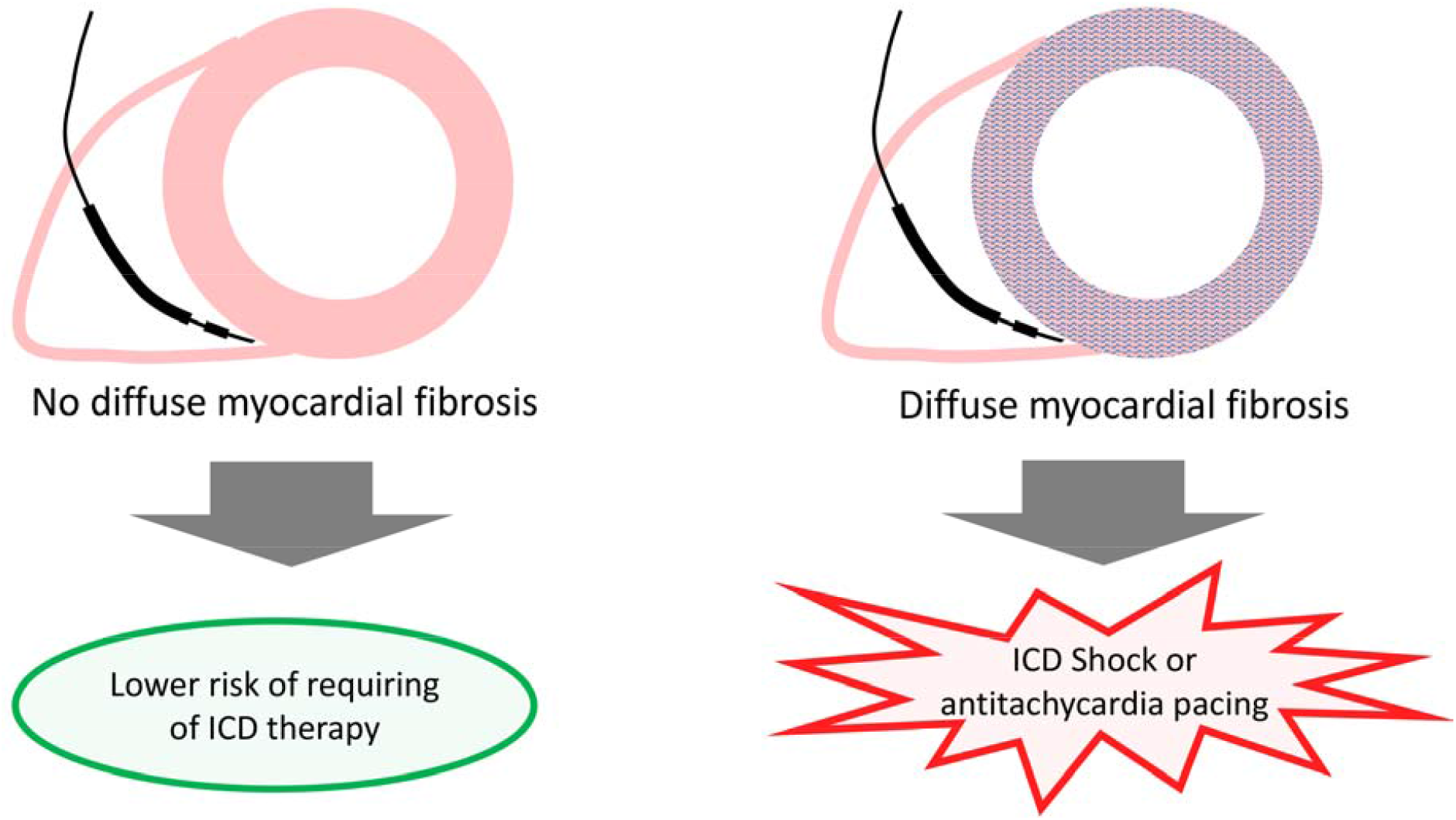
Diffuse myocardial fibrosis may represent a vulnerable phenotype and predispose to incident arrhythmia requiring therapy in implantable cardioverter defibrillator recipients.

**Supplemental Figure.**
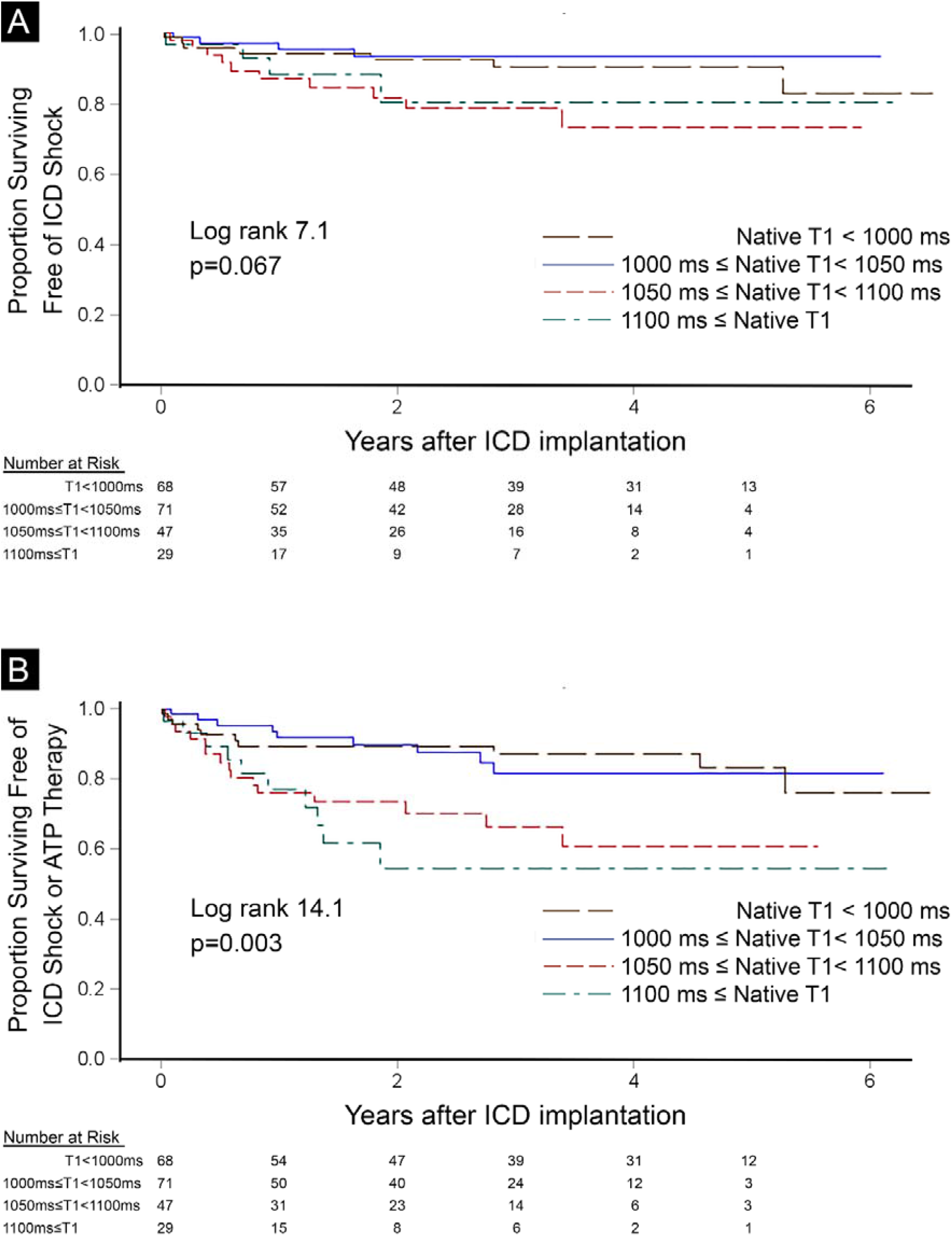
Native T1 categories did not associate with incident ICD shock (n=25, panel A) but did associate with the composite endpoint of ICD shock or ATP therapy (n=44, panel B) in 215 ICD recipients. These outcomes associations were not as robust compared to those of ECV in head to head univariable Cox regression model comparisons or in multivariable Cox regression models.

**Supplemental Table.** The overall prevalences of various LGE patterns in our cohort align with prevalences of various LGE patterns in prior reports.

| **Paper** | **Participants**<br>**(n)** | **Any LGE**<br>**prevalence** | **MI by**<br>**LGE**<br>**prevalence** | **Midwall**<br>**LGE** |
| --- | --- | --- | --- | --- |
| Gulati A, Jabbour A, Ismail TF et al. Association of fibrosis with mortality and sudden cardiac death in patients with nonischemic dilated cardiomyopathy. JAMA 2013;309:896-908.(15) | 472 | - | - | 30% |
| Pontone G, Guaricci AI, Andreini D et al. Prognostic Benefit of Cardiac Magnetic Resonance Over Transthoracic Echocardiography for the Assessment of Ischemic and Nonischemic Dilated Cardiomyopathy Patients Referred for the Evaluation of Primary Prevention Implantable Cardioverter-Defibrillator Therapy. Circ Cardiovasc Imaging 2016;9. (2) | 409 | 60% | 52% | Not reported |
| Gao P, Yee R, Gula L et al. Prediction of arrhythmic events in ischemic and dilated cardiomyopathy patients referred for implantable cardiac defibrillator: evaluation of multiple scar quantification measures for late gadolinium enhancement magnetic resonance imaging. Circ Cardiovasc Imaging 2012;5:448-56. (23) | 124 | 85% | Not reported | Not reported |
| Klem I, Weinsaft JW, Bahnson TD et al. Assessment of myocardial scarring improves risk stratification in patients evaluated for cardiac defibrillator implantation. J Am Coll Cardiol 2012;60:408-20. (17) | 137 | 78% | Not reported | Not reported |
| Current manuscript: Olausson E, Wertz J, Fridman Y, et al. Diffuse myocardial fibrosis associates with incident ventricular arrhythmia in implantable cardioverter defibrillator recipients. JACC Cardiovasc Imaging 2020 | **215** | **73%** | **46%** | **28%** |

## Notes

**Funding:** Dr. Miller is funded by a Clinician Scientist Award (CS-2015-15-003) from the National Institute for Health Research, Manchester, UK. Dr. Wong was supported by American Heart Association Scientist Development grant (Dallas, TX) and Children’s Cardiomyopathy Foundation grant. Dr. Schelbert was supported by a grant from The Pittsburgh Foundation (PA), Grant M2009□0068 to EBS; and an American Heart Association (Dallas, TX) Scientist Development grant (09SDG2180083) including a T. Franklin Williams Scholarship Award; funding provided by: Atlantic Philanthropies, Inc, the John A. Hartford Foundation, the Association of Specialty Professors, and the American Heart Association (Dallas, TX). Dr. Ugander was supported in part by grants from New South Wales Health, Heart Research Australia, and the University of Sydney. This work was also supported by Grant Number UL1-TR-001857 from the National Center for Research Resources (NCRR), a component of the National Institutes of Health (NIH), and NIH Roadmap for Medical Research (Bethesda, MD).

**Conflict of Interest Disclosures:** Dr. Miller received research support from Roche and Guerbet and serves as Scientific Advisor to Haya Therapeutics. Dr. Schelbert has accepted contrast material from Bracco Diagnostics for research purposes and serves as Scientific Advisor to Haya Therapeutics. Dr. Ugander is principal investigator on a research and development agreement regarding CMR between Siemens and Karolinska University Hospital. Dr. Saba has received research support from Boston Scientific. Dr. Cavalcante has received consulting fees from Boston Scientific, Medtronic and Abbott Vascular; and research grant support from Abbott Vascular, Edwards Lifesciences, Medtronic, Boston Scientific, Circle Cardiovascular Imaging and Medis. The remaining authors have nothing to disclose.

### Competing Interest Statement

Dr. Miller received research support from Roche and Guerbet and serves as Scientific Advisor to Haya Therapeutics. Dr. Schelbert has accepted contrast material from Bracco Diagnostics for research purposes and serves as Scientific Advisor to Haya Therapeutics. Dr. Ugander is principal investigator on a research and development agreement regarding CMR between Siemens and Karolinska University Hospital. Dr. Saba has received research support from Boston Scientific. Dr. Cavalcante has received consulting fees from Boston Scientific, Medtronic and Abbott Vascular; and research grant support from Abbott Vascular, Edwards Lifesciences, Medtronic, Boston Scientific, Circle Cardiovascular Imaging and Medis. The remaining authors have nothing to disclose.

### Funding Statement

Dr. Miller is funded by a Clinician Scientist Award (CS-2015-15-003) from the National Institute for Health Research, Manchester, UK. Dr. Wong was supported by American Heart Association Scientist Development grant (Dallas, TX) and Childrens Cardiomyopathy Foundation grant. Dr. Schelbert was supported by a grant from The Pittsburgh Foundation (PA), Grant M2009‐0068 to EBS; and an American Heart Association (Dallas, TX) Scientist Development grant (09SDG2180083) including a T. Franklin Williams Scholarship Award; funding provided by: Atlantic Philanthropies, Inc, the John A. Hartford Foundation, the Association of Specialty Professors, and the American Heart Association (Dallas, TX). Dr. Ugander was supported in part by grants from New South Wales Health, Heart Research Australia, and the University of Sydney. This work was also supported by Grant Number UL1-TR-001857 from the National Center for Research Resources (NCRR), a component of the National Institutes of Health (NIH), and NIH Roadmap for Medical Research (Bethesda, MD).

### Author Declarations

The Institutional Review Board at the University of Pittsburgh Medical Center approved the study, and all subjects provided written informed consent.

## References

1. Schelbert EB, Piehler KM, Zareba KM et al. Myocardial Fibrosis Quantified by Extracellular Volume Is Associated With Subsequent Hospitalization for Heart Failure, Death, or Both Across the Spectrum of Ejection Fraction and Heart Failure Stage. J Am Heart Assoc 2015;4:e002613.

2. Treibel TA, Fridman Y, Bering P et al. Extracellular volume associates with outcomes more strongly than native or post-contrast myocardial T1. JACC Cardiovasc Imaging 2019;[Epub ahead of print]:.

3. Yang EY, Ghosn MG, Khan MA et al. Myocardial Extracellular Volume Fraction Adds Prognostic Information Beyond Myocardial Replacement Fibrosis. Circ Cardiovasc Imaging 2019;12:e009535.

4. Fröjdh F, Fridman Y, Bering P et al. Extracellular volume and global longitudinal strain both associate with outcomes but correlate minimally JACC Cardiovasc Imaging 2020;in press.

5. Wong TC, Piehler K, Meier CG et al. Association between extracellular matrix expansion quantified by cardiovascular magnetic resonance and short-term mortality. Circulation 2012;126:1206–16.

6. Chin CW, Everett RJ, Kwiecinski J et al. Myocardial Fibrosis and Cardiac Decompensation in Aortic Stenosis. JACC Cardiovasc Imaging 2016;10:1320–1333.

7. Duca F, Zotter-Tufaro C, Kammerlander AA et al. Cardiac extracellular matrix is associated with adverse outcome in patients with chronic heart failure. Eur J Heart Fail 2016;Epub ahead of print.

8. Duca F, Kammerlander AA, Zotter-Tufaro C et al. Interstitial Fibrosis, Functional Status, and Outcomes in Heart Failure With Preserved Ejection Fraction: Insights From a Prospective Cardiac Magnetic Resonance Imaging Study. Circ Cardiovasc Imaging 2016;9:e005277.

9. Schelbert EB, Fridman Y, Wong TC et al. Temporal relation between myocardial fibrosis and heart failure with preserved ejection fraction: association with baseline disease severity and subsequent outcome JAMA Cardiol 2017;2:1–12.

10. Wong TC, Piehler K, Kang IA et al. Myocardial Extracellular Volume Fraction Quantified By Cardiovascular Magnetic Resonance is Increased in Diabetes and Associated with Mortality and Incident Heart Failure Admission. Eur Heart J 2014;35:657–664.

11. Youn JC, Hong YJ, Lee HJ et al. Contrast-enhanced T1 mapping-based extracellular volume fraction independently predicts clinical outcome in patients with non-ischemic dilated cardiomyopathy: a prospective cohort study. Eur Radiol 2017;27:3924–3933.

12. Kanagala P, Cheng ASH, Singh A et al. Relationship Between Focal and Diffuse Fibrosis Assessed by CMR and Clinical Outcomes in Heart Failure With Preserved Ejection Fraction. JACC Cardiovasc Imaging 2019.

13. Santini M, Lavalle C, Ricci RP. Primary and secondary prevention of sudden cardiac death: who should get an ICD? Heart 2007;93:1478–83.

14. Schelbert EB, Sabbah HN, Butler J, Gheorghiade M. Employing Extracellular Volume Cardiovascular Magnetic Resonance Measures of Myocardial Fibrosis to Foster Novel Therapeutics. Circ Cardiovasc Imaging 2017;10:e005619.

15. Gulati A, Jabbour A, Ismail TF et al. Association of fibrosis with mortality and sudden cardiac death in patients with nonischemic dilated cardiomyopathy. JAMA 2013;309:896–908.

16. Halliday BP, Gulati A, Ali A et al. Association Between Midwall Late Gadolinium Enhancement and Sudden Cardiac Death in Patients With Dilated Cardiomyopathy and Mild and Moderate Left Ventricular Systolic Dysfunction. Circulation 2017;135:2106–2115.

17. Klem I, Weinsaft JW, Bahnson TD et al. Assessment of myocardial scarring improves risk stratification in patients evaluated for cardiac defibrillator implantation. J Am Coll Cardiol 2012;60:408–20.

18. Almehmadi F, Joncas SX, Nevis I et al. Prevalence of myocardial fibrosis patterns in patients with systolic dysfunction: prognostic significance for the prediction of sudden cardiac arrest or appropriate implantable cardiac defibrillator therapy. Circ Cardiovasc Imaging 2014;7:593–600.

19. Leyva F, Zegard A, Acquaye E et al. Outcomes of Cardiac Resynchronization Therapy With or Without Defibrillation in Patients With Nonischemic Cardiomyopathy. J Am Coll Cardiol 2017;70:1216–1227.

20. Gutman SJ, Costello BT, Papapostolou S et al. Reduction in mortality from implantable cardioverter-defibrillators in non-ischaemic cardiomyopathy patients is dependent on the presence of left ventricular scar. Eur Heart J 2019;40:542–550.

21. Di Marco A, Anguera I, Schmitt M et al. Late Gadolinium Enhancement and the Risk for Ventricular Arrhythmias or Sudden Death in Dilated Cardiomyopathy: Systematic Review and Meta-Analysis. JACC Heart Fail 2017;5:28–38.

22. Pontone G, Guaricci AI, Andreini D et al. Prognostic Benefit of Cardiac Magnetic Resonance Over Transthoracic Echocardiography for the Assessment of Ischemic and Nonischemic Dilated Cardiomyopathy Patients Referred for the Evaluation of Primary Prevention Implantable Cardioverter-Defibrillator Therapy. Circ Cardiovasc Imaging 2016;9.

23. Gao P, Yee R, Gula L et al. Prediction of arrhythmic events in ischemic and dilated cardiomyopathy patients referred for implantable cardiac defibrillator: evaluation of multiple scar quantification measures for late gadolinium enhancement magnetic resonance imaging. Circ Cardiovasc Imaging 2012;5:448–56.

24. Voskoboinik A, Wong MCG, Elliott JK et al. Absence of late gadolinium enhancement on cardiac magnetic resonance imaging in ventricular fibrillation and nonischemic cardiomyopathy. Pacing Clin Electrophysiol 2018.

25. Tamarappoo BK, John BT, Reinier K et al. Vulnerable myocardial interstitium in patients with isolated left ventricular hypertrophy and sudden cardiac death: a postmortem histological evaluation. J Am Heart Assoc 2012;1:e001511.

26. Beltrami CA, Finato N, Rocco M et al. Structural basis of end-stage failure in ischemic cardiomyopathy in humans. Circulation 1994;89:151–63.

27. Beltrami CA, Finato N, Rocco M et al. The cellular basis of dilated cardiomyopathy in humans. J Mol Cell Cardiol 1995;27:291–305.

28. Lewis GA, Dodd S, Clayton D et al. Pirfenidone in heart failure with preserved ejection fraction: a randomized phase 2 trial. Nat Med 2021;27:1477–1482.

29. Masci PG, Schuurman R, Andrea B et al. Myocardial fibrosis as a key determinant of left ventricular remodeling in idiopathic dilated cardiomyopathy: a contrast-enhanced cardiovascular magnetic study. Circ Cardiovasc Imaging 2013;6:790–9.

30. Kim RJ, Albert TS, Wible JH et al. Performance of delayed-enhancement magnetic resonance imaging with gadoversetamide contrast for the detection and assessment of myocardial infarction: an international, multicenter, double-blinded, randomized trial. Circulation 2008;117:629–37.

31. Schelbert EB, Butler J, Diez J. Why Clinicians Should Care About the Cardiac Interstitium. JACC Cardiovasc Imaging 2019.

32. Felker GM, Shaw LK, O’Connor CM. A standardized definition of ischemic cardiomyopathy for use in clinical research. J Am Coll Cardiol 2002;39:210–8.

33. Wong TC, Piehler KM, Zareba KM et al. Myocardial damage detected by late gadolinium enhancement cardiovascular magnetic resonance is associated with subsequent hospitalization for heart failure. J Am Heart Assoc 2013;2:e000416.

34. Piehler KM, Wong TC, Puntil KS et al. Free-breathing, motion-corrected late gadolinium enhancement is robust and extends risk stratification to vulnerable patients. Circ Cardiovasc Imaging 2013;6:423–32.

35. Schelbert EB, Testa SM, Meier CG et al. Myocardial extravascular extracellular volume fraction measurement by gadolinium cardiovascular magnetic resonance in humans: slow infusion versus bolus. J Cardiovasc Magn Reson 2011;13:16.

36. Moon JC, Messroghli DR, Kellman P et al. Myocardial T1 mapping and extracellular volume quantification: a Society for Cardiovascular Magnetic Resonance (SCMR) and CMR Working Group of the European Society of Cardiology consensus statement. J Cardiovasc Magn Reson 2013;15:92.

37. Arheden H, Saeed M, Higgins CB et al. Measurement of the distribution volume of gadopentetate dimeglumine at echo-planar MR imaging to quantify myocardial infarction: comparison with 99mTc-DTPA autoradiography in rats. Radiology 1999;211:698–708.

38. Schelbert EB, Piehler KM, Zareba KM et al. Myocardial Fibrosis Quantified by Extracellular Volume Is Associated With Subsequent Hospitalization for Heart Failure, Death, or Both Across the Spectrum of Ejection Fraction and Heart Failure Stage. J Am Heart Assoc 2015;4:e002613.

39. Lam B, Stromp TA, Hui Z, Vandsburger M. Myocardial native-T1 times are elevated as a function of hypertrophy, HbA1c, and heart rate in diabetic adults without diffuse fibrosis. Magn Reson Imaging 2019;61:83–89.

40. Hamon D, Algalarrondo V, Gandjbakhch E et al. Outcome and incidence of appropriate implantable cardioverter-defibrillator therapy in patients with cardiac amyloidosis. Int J Cardiol 2016;222:562–8.

41. Di Marco A, Brown PF, Bradley J et al. Extracellular volume fraction improves risk-stratification for ventricular arrhythmias and sudden death in non-ischaemic cardiomyopathy. Eur Heart J Cardiovasc Imaging 2022.

42. Claridge S, Mennuni S, Jackson T et al. Substrate-dependent risk stratification for implantable cardioverter defibrillator therapies using cardiac magnetic resonance imaging: The importance of T1 mapping in nonischemic patients. J Cardiovasc Electrophysiol 2017;28:785–795.

43. Chen Z, Sohal M, Voigt T et al. Myocardial tissue characterization by cardiac magnetic resonance imaging using T1 mapping predicts ventricular arrhythmia in ischemic and non-ischemic cardiomyopathy patients with implantable cardioverter-defibrillators. Heart Rhythm 2015;12:792–801.

44. Xue H, Greiser A, Zuehlsdorff S et al. Phase-sensitive inversion recovery for myocardial T1 mapping with motion correction and parametric fitting. Magn Reson Med 2013;69:1408–20.

45. Xue H, Shah S, Greiser A et al. Motion correction for myocardial T1 mapping using image registration with synthetic image estimation. Magn Reson Med 2012;67:1644–1655.

46. Kober L, Thune JJ, Nielsen JC et al. Defibrillator Implantation in Patients with Nonischemic Systolic Heart Failure. N Engl J Med 2016;375:1221–30.

47. Selvanayagam JB, Hartshorne T, Billot L et al. Cardiovascular magnetic resonance-GUIDEd management of mild to moderate left ventricular systolic dysfunction (CMR GUIDE): Study protocol for a randomized controlled trial. Ann Noninvasive Electrocardiol 2017;22.

48. Ramires FJ, Mansur A, Coelho O et al. Effect of spironolactone on ventricular arrhythmias in congestive heart failure secondary to idiopathic dilated or to ischemic cardiomyopathy. Am J Cardiol 2000;85:1207–11.

49. Guerra F, Malagoli A, Contadini D et al. Global Longitudinal Strain as a Predictor of First and Subsequent Arrhythmic Events in Remotely Monitored ICD Patients With Structural Heart Disease. JACC Cardiovasc Imaging 2020;13:1–9.

50. Halliday BP, Baksi AJ, Gulati A et al. Outcome in Dilated Cardiomyopathy Related to the Extent, Location, and Pattern of Late Gadolinium Enhancement. JACC Cardiovasc Imaging 2019;12:1645–1655.

